# An Empty Scoping Review of Emergency Department to School Transition for Youth with Mental Health Concerns

**DOI:** 10.1101/2024.01.23.24301673

**Authors:** Lora Henderson Smith, Emily Warren, Natalie Hendrickson, Kate Joshua

## Abstract

The number of youth Emergency Department (ED) visits due to mental health concerns has been steadily increasing with a large number of youth being referred from school. Despite the increase in ED visits, there has not been an increase in the number of students who are actually admitted to the hospital. Further, youth referred from school are more likely to be discharged from the ED. Given the unique relationship between school and ED referrals and the large number of youth who do not require hospitalization, this study sought to understand how schools are supporting students who return to school after an ED visit. We conducted a scoping review to identify programs and practices to support ED to school transition. Two reviewers screened 907 manuscripts, but none of the manuscripts met the inclusion criteria. We discuss the importance of supporting students returning to school from the ED and draw from the literature on hospital to school transition to make recommendations for educators.

---

Pediatric emergency department (ED) visits due to mental health disorders have been on the rise with a 60% increase from 2007-2016 and a 329% increase in visits due to self-harm (Lo et al., 2020). This trend continued beyond 2016 and was exacerbated by the COVID-19 pandemic (Bommersbach et al., 2023; Yard et al., 2020). K-12 schools are a primary referral source for youth mental health ED visits (Soto et al., 2009) and there is a unique association between the school calendar and ED visits with mental health related ED visits decreasing during the summer months (Ali et al., 2012; Nixon et al., 2021; Copeland et al., 2022). Although many students are referred to the ED, the number of youth actually admitted to the hospital was found to be as low as 14% in one study (Gill et al., 2017). In a study of youth ages 5-18 referred to the ED from school due to agitation, youth were more likely to be Black, Latinx, or identified as a special education student (Toliver et al., 2022). Further, youth referred from school were more likely to be discharged home than youth referred from other sources (Toliver et al., 2022). Past studies have found that youth ages 15-18 who discharged from the ED were less likely to have care coordination services put in place (Lynch et al., 2021) putting these youth at further risk of returning to the ED and perhaps in greater need for school-based services and supports upon their return.

Services provided through school and school-based health centers can remove barriers to care that exist for some groups of students (Cutuli, 2022). Despite schools’ obligation to meet the educational needs of all students, school professionals report feeling unprepared to support students after psychiatric hospitalizations and this is likely true for ED visits as well (Tougas et al., 2019). Alternatively, there is little research on how ED providers are trained or how they conceptualize their role in supporting youth with mental health concerns serious enough to warrant an ED visit but who do not meet criteria for hospitalization. Given the dearth of literature on the ED to school transition, this scoping review examines existing practices and programs for supporting youth returning to school after an ED visit.

## Background on Youth Emergency Department Visits

There has been a significant increase in the number of youth visiting the ED with a mental health diagnosis compared to visits for other reasons (Cutler et al., 2019). The most common reason for a pediatric mental health ED visit is suicidality while other referral concerns include defiance, aggression, depression, anxiety, and psychosis (Bommersbach et al., 2023; Grudkinoff et al., 2015b). Suicidality is more commonly observed among females, individuals with depression, and those who indicate a precipitating peer conflict (2015b). The COVID-19 pandemic contributed to worsening adolescent mental health and in particular, there was a 51% increase in the number of girls requiring an ED visit after a suicide attempt (Yard et al., 2021). In a sample of 1,062 youth visiting a pediatric ED with psychiatric concerns, Black youth represented the greatest volume of patients (68.2%), followed by Latinx youth (28.9%) and White youth (2.5%) (Grudkinoff et al., 2015a). Underinsured youth and those with public insurance are also more likely to be seen in the ED (Hoge et al., 2022).

Notably, a large number of youth presenting to the ED are discharged without psychiatric hospitalization. However, youth who visit an ED at a hospital with an inpatient unit are more likely to be hospitalized (Cutler et al., 2019). Soto and colleagues (2009) found that 29% (n = 1,062) of youth patients presenting to a pediatric ED with psychiatric concerns required hospitalization. A larger sample of 118,851 youth had a much lower rate of hospitalization with only 14% of youth being admitted to the hospital after an ED visit. This is especially salient among Black preadolescents, with 93% of 504 patients ages 8-12 being discharged despite presenting with suicidal thoughts and/or behaviors (Vidal et al., 2023).

## The Relationship Between Schools and Emergency Department Visits

There is a unique relationship between school and ED visits with 44% of mental health emergency department visits resulting from school referrals (Grudkinoff et al., 2015a). School referrals may account for such a large percentage of the total volume of mental health ED visits due to school staff members’ lower threshold for labeling behavior as environmentally inappropriate as compared to parents or guardians (Goldstein et al., 2005). Furthermore, many school districts implement a zero-tolerance policy for violent behaviors and drug violations, both of which are cited as circumstances leading to an ED referral (Grupp-Phelan et al., 2009; Grudkinoff et al., 2015a).

Individuals presenting in the ED for suicidal ideation are more likely to be referred by a school than those presenting with a suicide attempt (Grudkinoff et al., 2015b). This may be because of mandated suicide risk assessments administered by school staff when concerns regarding self-injurious thoughts or behaviors are raised by teachers, peers, or the student themself (Crepeau-Hobson, 2013). According to Kodish et al. (2020), school staff describe this assessment as necessary to ensure a student’s safety, while students view the process as disciplinary despite being assured otherwise. Students also report perceptions that the risk assessment has a low threshold for moderate to high-risk profiles (Kodish et al., 2020). Research by Grudkinoff et al. (2015b) supports this idea of a lower threshold, suggesting that many school referrals for suicidality may be inappropriate for ED-level intervention. Yet, Grudkinoff et al. (2015a) report that 82% of 200 children referred by their school were not evaluated by a school mental health professional or nurse prior to arriving at the ED. Further, communication between school staff and ED clinicians is insufficient contributing to discontinuity in care (Alvarado et al., 2020). Students and parents alike express feelings of support from the school in response to risk detection; thus, direct referral to the ED without prior evaluation may leave families feeling unsupported by the school in the ED experience (Kodish et al., 2020).

For all cases of suicidality, school referrals were associated with a greater likelihood of discharge without psychiatric hospitalization (Grudkinoff et al., 2015b). Discharge without follow-up is more common among individuals expressing suicidality as compared to those visiting the ED for other mental health concerns (Grudkinoff et al., 2015a). Among all ED visits, only a small percentage (7.8%) of children and adolescents referred to the ED by their school were admitted to a psychiatric hospital; most were discharged without a follow-up appointment (Grudkinoff et al., 2015a).

Considering that the risk of a second ED visit is highest in the month post-discharge, it is essential for school mental health professionals to take an active role in supporting students as they return to school following an ED visit (King et al., 2019). This may be especially important for individuals presenting with suicidality, as the majority are discharged home from the ED without a scheduled follow-up (Grudkinoff et al., 2015b). Research by Goldstein et al. (2005) suggests that school may increase stress and mental illness symptom severity among children and adolescents, emphasizing the necessity of a school reintegration plan to ensure return to an environment laden with academic, counseling, and behavioral interventions.

## Interventions to Support Youth in the Emergency Department

Several interventions have been used to support youth in the ED and promote positive mental health outcomes (Asarnow et al., 2011; Donaldson et al., 1997; Kodish et al., 2023; Rengasamy & Sparks, 2019; Ryan et al., 2022). Donaldson and colleagues (1997) implemented an intervention to increase the likelihood of patients seeking follow up services. Youth and their caregiver completed a psychotherapy compliance enhancement intervention in the ED and then youth in the experimental condition (n=28) received three follow-up phone calls focused on problem-solving and suicidal ideation. The youth in the experimental condition were more likely to attend follow up appointments and had no more suicide attempts compared to 9% of youth in the control group (n= 78) who made a subsequent attempt. In another follow-up phone call intervention, youth receiving a multi-call intervention (n = 72) consisting of suicide assessment, safety planning, medication and weapon safety discussions demonstrated a lower rate of suicidal behavior and greater confidence in their safety plan than youth receiving on one follow up phone call (n = 70) (Renegasmy & Sparks, 2019). In addition to telephone-based interventions, one study using text messages to follow up with adolescents after ED visits due to suicidal thoughts or behavior was found to be acceptable and feasible with youth reporting decreased suicidal thoughts and behaviors after receiving the texts (Ryan et al., 2022).

Other interventions have been delivered directly in the ED. For example, the Family Intervention for Suicide Prevention consisted of ED clinician training, a family crisis therapy session, and follow up contacts to facilitate linkage to care. This intervention was adapted specifically for racial and ethnic minority youth and named Safe Alternatives for Teens and Youths–Acute (SAFETY-A) (Kodish et al., 2023). Youth receiving the treatment were more likely to engage in follow up treatment after the ED visit. Although there are phone and ED-based intervention, there is a need for interventions to support the return to school.

## School Supports for Students After Hospitalization

Although the research on supporting students after an ED visit is limited, there is some literature on supporting students after a medical or psychiatric hospitalization.

### Medical Hospitalization

Like children experiencing mental health crises, extant research on supportive transition for youth returning to school following a hospital visit for physical health conditions such as chronic illnesses (e.g. asthma, blood disorders, cancer, cardiac conditions, cystic fibrosis, diabetes, and juvenile idiopathic arthritis) or traumatic brain injuries suggests that these students experience academic and social difficulties upon their return (Ireys, 2001; Glang et al., 2018; Wikel and Markelz, 2023). While the structure of school reentry programs for these youth is highly variable, having a hospital to school transition program in place is related to fewer absences and higher self-reported feelings of social connection at school upon their return (Wikel & Markelz, 2023).

Successful reintegration has been associated with the creation of a multidisciplinary team of medical professionals, school personnel, and the student’s family to determine accommodations tailored to the child’s individual needs (Nabors et al., 2005; Shaw & McCabe, 2008). Given that school personnel frequently report feeling unqualified to accommodate the academic and social needs of youth experiencing chronic illnesses, this approach may mitigate such concerns (Wikel & Marklez, 2023). A similar approach to school reentry involves a hospital liaison tasked with facilitating communication between hospitals, schools, and families (Kaffenberger, 2006; Moore et al., 2009; Wikel & Markelz, 2023). One such example is the STEP model of school reentry to support students following hospitalization for a traumatic brain injury, as described by Glang and colleagues (2018). In this model, hospitals contact a Department of Education liaison who then assigns a trained transition (STEP) facilitator to the child’s case. The STEP facilitator’s role is to inform the child’s school of their hospitalization and condition, consult with hospital staff and school personnel to ensure that necessary academic and social accommodations were provided in the school environment, and track the student’s progress upon their return to school to inform modification. STEP was not associated with significant differences in behavioral and executive functioning compared to the control group at one-month post-discharge and one-year follow-up points (Glang et al., 2018); yet, the use of hospital liaisons in school reentry may reconcile discrepancies between hospital staff lacking knowledge of the education system and school personnel with limited knowledge about their students’ medical conditions (Wikel & Markelz, 2023).

### Psychiatric Hospitalization

Those attempting to support youth returning to school from the ED may be able to learn from the literature on supporting students returning to school after a psychiatric hospitalization. These transitions typically occur when an adolescent has been hospitalized in a short-term psychiatric program and discharged to the community with the intention to return to school. These hospitalizations can last anywhere from days to months, and during this time, a student may be unenrolled from their home school district and enrolled in the hospital education program. These programs typically offer onsite classes, and for students that are eligible for special education, they should also be adhering to the services outlined in the child’s Individualized Education Program (DBHDS, 2023). When the hospitalization is expected to be short-term, some youth may not receive any educational programming. This makes it challenging to create universal standards for hospital to school transitions, as each case may have different circumstances. Despite this importance policies and procedures for school reintegration, 45% of school districts report having no formal re-entry protocol, and less than a third of districts reported formal, written protocols (Marraccini et al., 2019).

The formal or informal transition process usually includes a meeting with a school counselor or administrator upon return to the building to determine additional support needs and develop a school re-entry plan. The Behavioral Health Integrated Resources for Children project (BIRCh) developed a hospital transition protocol that incorporates working with the school, family, hospital, and student to develop a plan for their return to school (BIRCh, 2021). The protocol includes a timeline that begins at the point of admission, so schools may address reintegration needs as soon as possible, and continues throughout the student’s transition back to school (Tougas et al., 2022). Each plan is individualized to the student’s particular needs with school staff partnering with the student and their family to identify safety, academic, and social emotional support needs (Preyde et al., 2017). Some options include having the student return to a partial-day schedule, having regularly scheduled check-in/check-out meetings with the counselor, or participation in psychoeducational or coping groups (Marraccini et al., 2022). These activities take coordination, requiring schools and families communicate prior to discharge so the school may make the necessary adjustments.

At the same time as coordinating school services for discharge, hospital staff are working to connect the child with follow-up mental health services in their community (James et al., 2010). Types of aftercare services include intensive mental health services (e.g., partial hospitalization, intensive outpatient, residential treatment), non-intensive mental health services (e.g., outpatient therapy, anger management, home-based counseling), and other services (e.g., case management, AA/NA support groups, mentoring, involvement with child welfare) (James et al., 2010). Some children may be connected to mental health services prior to hospitalization, so they may focus on scheduling their next appointment versus scheduling an intake to a new service. Individuals are more likely to attend outpatient appointments if they have been scheduled prior to discharge (Smith et al., 2020). It may be necessary to work with families to explore potential resources and anticipated barriers to continuing recommended treatment services (Gulliver et al., 2010).

In a comprehensive review of school reintegration after psychiatric hospitalization programs, Tougas et al. (2022) concluded that although these programs are rare and difficult to implement, there are four identified programs out of eight total that have promising results: the Bridge program, the Bridges program, the Bridge for Resilient Youth in Transition (BRYT), and the School Transition Program (STP). These programs were promising based on their inclusion of multidisciplinary team members, a multicomponent intervention, formal transition plan, gradual reintegration, and continued follow-up support (Tougas et al., 2022). A limitation of this previous research is that these programs are generally local initiatives with small sample sizes, and more robust research is needed; however, promising practices in hospital to school transition may be able to inform successful re-entry plans for students after ED visits as well.

### Emergency Department Visits

When children are sent to the ED and released, they may not trigger the same coordination of school services as a student who required a hospitalization, and they possibly have not even missed a day of class. For students who were not referred to the ED by school, school officials may count on the student’s family to notify them of the ED visit (Tougas et al., 2019; Clemens et al., 2011). While schools may have accommodations in place for physically injured children returning to school, such as access to an elevator, change in classroom seating, extended hallway transition time, there are not universally implemented accommodations or resources available to students discharged from EDs for psychological distress. Although these students who visit the ED may not end up placed in a psychiatric hospitalization program, this does not mean they are not in crisis (Spirito & Esposito-Smythers, 2006). They may still have significant mental health concerns and troubling behaviors, and they could benefit from those same accommodations that are offered to students transitioning from psychiatric hospital programs (Rotheram-Borus et al., 2000).

## Theoretical Framework

Schools play a vital role in supporting students’ mental health and this is particularly true for students from marginalized and minoritized backgrounds (Hoover & Bostic, 2020). As such, this study is guided by the Interconnected Systems Framework, which supports the integration of school mental health services and Positive Behavioral Interventions and Supports (Eber et al., 2019; Weist et al., 2022). Specifically, the ISF leverages community resources and aims to provide one system of service delivery for education and mental health related services within a multi-tiered system (Eber et al., 2019). Within this framework, there is a District-Community Leadership Team that consists of key community members such as education and mental health leaders as well as leaders from other systems of care, family members, and students (Barret et al., 2013). At the school level, the MTSS team within a school using the Interconnected Systems Framework, includes mental health clinicians in addition to the general education, special education, administrators, and other interventionists and individuals who typically comprise the team (Barret et al., 2013). Within an Interconnected Systems Framework, school mental health clinicians are well positioned to support the mental health needs of students who need a Tier 3 level of social and emotional support such as those returning to school after an ED visit. Further, the integration between systems (i.e., schools, mental health, community resources) is ideal for supporting student mental health across texts.

## Current Study

Given that the majority of youth who visit the ED due to mental health concerns do not require an inpatient psychiatric hospitalization (Gill et al., 2017; Grupp-Phelan et al., 2009) and that over one third of students are referred to the ED from school (Soto et al., 2009) and will subsequently return to school, it is important to identify best practices in supporting the transition back to school after an ED visit. As such, we conducted a scoping review to answer the following research question: What is the existing literature base for ED to school transition for K-12 students?

## Methods

Given the limited literature base on supporting students after ED visits, we conducted a scoping review, which is a research method that can be used to broadly summarize literature. Our scoping review and data analysis methods were guided by the Preferred Reporting Items for Systematic Reviews-Extension for Scoping Reviews (Tricco et al., 2018). The search strategies were developed by the research team with guidance from a librarian with expertise in conducting scoping reviews as well as health sciences research. The strategies were run in education and health sciences databases including Education Research Complete, PsycInfo, PubMed/MEDLINE, and Web of Science.

For the inclusion criteria, we were specifically interested in any hospital, school, or community provider practices that have been used to support K-12 students who visited the ED due to mental health concerns and were discharged home.

### Search Strategy

Given that each manuscript must describe an intervention or specific practice(s) used to support students returning to school after a mental health ED visit, each had at least one related term/phrase from the following categories:

- **Population age of study:** pediatric, child, youth, student, teenager, adolescent, kid
- **Setting:** emergency department AND school (primary, secondary, elementary, junior high, middle, high school)
- **Mental Health:** mental health, behavioral health
- **Concept of transition between settings:** “Return to,” transition, reentry, discharge When available, terms were translated into respective database terminologies and additional keywords were used when necessary and applicable.

Each manuscript was reviewed by two trained members of the research team. Any conflicts were discussed by the research team until consensus was reached.

**Figure 1.**
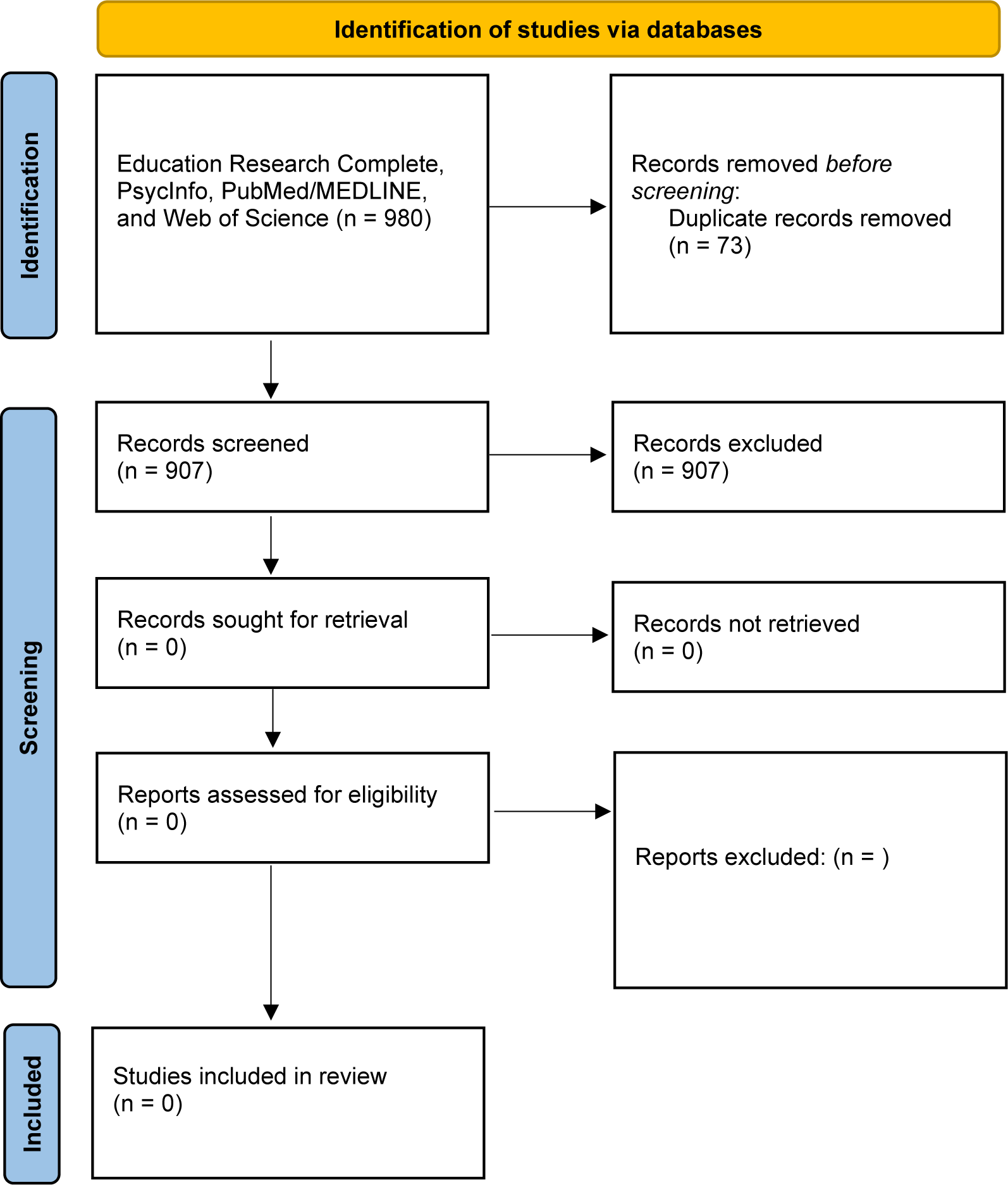
PRISMA 2020 Flow Diagram for Screening.

Figure template from (Page et al., 2021)

### Compliance with Ethical Standards

The authors do not have any financial or non-financial conflicts of interest. This study did not require approval by an Institutional Review Board or any consenting procedures given that it solely entailed a review of the existing literature base.

## Results

Our search strategies resulted in the identification of 907 manuscripts that were reviewed by two members of the research team. Of these 907 manuscripts, none of them provided guidance or highlighted programs or practices to support the transition of K-12 youth returning to school after an ED visit.

## Discussion

Reviews, such as ours, yielding no eligible studies are often referred to as “empty reviews” (Lang et al., 2007; Yaffe et al., 2012). Initially, it may seem that there is nothing to be reported from an empty review; however, these reviews can serve several purposes. First, empty reviews can reveal who the researchers are who are interested in the topics at hand (Lang et al., 2007). Further, they still serve the purpose of summarizing the literature base or in other words, identifying significant games in the literature (Lang et al., 2007). As such, our study indicates the need for an increased focus on supporting students returning to school after ED visits due to mental health concerns.

Studies indicate a high percentage of youth who visit the ED are discharged back home and often without a follow-up appointment scheduled. In one retrospective chart review including 452 pediatric patients (20 or younger) from four urban EDs, 52% of youth were admitted to the hospital (Grupp-Phelan et al., 2009). On the other hand, a larger study of 118,851 10 to 24-year-olds found that only 14% of youth who visited the ED were admitted (Gill et al., 2017). In considering the youth who visit the ED, about one third of them are referred from school (Soto et al., 2009). Given the sizable number of youth who will be referred to the ED by school staff and then discharged to return to their regular routine and school attendance, school staff must be prepared to support these children who may still be experiencing significant mental health concerns.

Meeting the needs of youth returning to school after an ED visit is particularly challenging given their various needs. Some youth may be appropriately discharged because they do not require a higher level of care; however, others are sometimes discharged because there are no inpatient beds available to them or for other systemic reasons. Although the majority of youth seen in the ED with a primary mental health diagnosis are experiencing suicidal ideation or have made an attempt, other reasons for visits include aggression, psychosis, anxiety, and disordered eating just to name a few. Given the variation of presenting concerns, schools face the challenge of needing to develop individualized plans to support each students’ personal needs as they return to school.

School and ED-based clinicians can draw from the hospital to school transition literature as they work to identify best practices in supporting students returning to school after an ED visit. Consistent with findings from this review, our ongoing conversations with school-based practitioners suggest that most schools and districts are not implementing formal ED to school transition protocols. However, 16% of school psychologists report having formal hospital to school transition protocols and 45% report informal procedures (Marraccini et al., 2019). Hospital to school re-entry protocols could be used or adapted for ED transition as well.

There are also equity issues to consider related to youth ED visits for mental health concerns. Black, Latinx, and publicly-insured youth are overrepresented in the number of youth who visit the ED with a mental health condition (Hoge et al., 2022). These same youth are also more likely to receive mental health services in schools (Ali et al., 2019; Wilk et al., 2022). ED and hospital staff must consider how social determinants of health impact this marginalized and minoritized groups and how they can best partner to support the transition back to school. Interventions in the ED have sought to facilitate linkage to care. For example, the Family Intervention for Suicide Prevention (FISP) was a single session delivered in the ED that is based on cognitive behavioral theories and seeks to increase the likelihood of adolescents receiving ongoing services after discharge (Hughes & Asarnow et al., 2015). FISP is now known as Safe Alternatives for Teens and Youths–Acute (SAFETY-A) and has recently been delivered in the ED to specifically promote continuity in care for youth from racial and ethnic minority backgrounds (Kodish et al., 2023). SAFETY-A is currently being studied for supporting youth in school who are experiencing mental health crises (Yu et al., 2023).

Ideally, schools can serve as sites of prevention, but some kids will inevitably require Tier 2 and Tier 3 mental health supports in school. Schools that can meet the needs of students after ED visits play an important role in preventing future ED visits and possible hospitalizations. Students who feel support and connected at school are less likely to return to the ED indicating the importance of engaging in best practices to support student mental health.

### Recommendations for ED to School Transition

The Interconnected Systems Framework offers an ideal approach to meeting the needs of students with mental health challenges such as those returning to school after an ED visit. Given the integration of community resources with education and mental health related services within a multi-tiered system, continuity of care could easily be facilitated for these students (Eber et al., 2019). Further, they would have hopefully already engaged with school mental health professionals for Tier 1 and Tier 2 interventions allowing trust and rapport to already be in place when Tier 3 interventions are needed after an ED visit. The Interconnected Systems Framework also promotes collaboration across systems which can improve relationships between schools and EDs to facilitate better case for students who must navigate each system.

Some recommendations for hospital to school transition can be adapted to support students returning to school after an ED visits. Some of these practices included: information sharing between the ED, school, and family, holding a school re-entry meeting, creating a school re-entry plan, and safety planning (Marraccini et al., 2022; Savina et al., 2014; Tougas et al., 2019). School re-entry after an ED visit will likely focus more on social and emotional support given that there may be less make-up work to be completed than after a hospitalization. However, there may be times when a student has been struggling with mental health and despite being present at school, there is a need to identify and support their academic needs. After an ED visit, it can be particularly helpful if school mental health professionals or administrators are able to share resource maps or community resources to help facilitate linkages to care that are not always put in place during a short ED visit. Lastly, beyond the initial school re-entry meeting, school staff should follow up with the student and family regularly to help support their transition back to school and hopefully minimize risk of readmission.

### Limitations

Our search did not result in the inclusion of any ED to school transition programs or practices although it does provide insightful information on the state of the field. However, there are ED-based interventions that are designed to support continuity of care. While reviewing these types of programs was beyond the scope of our review as we were specifically interested in understanding how schools can support students, these programs may provide some insight into what practices schools can incorporate into their protocols for students returning to school after an ED visit.

### Conclusion

Given the steady increase in youth visiting the ED with mental health concerns and the unique relationship between schools and ED visits, it is imperative that ED clinicians and school personnel collaborate to meet students’ needs. This review demonstrates a clear need for researchers and practitioners to identify best practices for supporting youth returning to school. While robust school mental health services after an ED visit are needed to support students who may still be in crisis, they can also serve a vital role in prevention and triaging as well. Although the literature on ED to school transition is limited, the hospital to school transition literature provides a starting point for supporting youth after ED visits.

## Data Availability

All data produced in the present study are available upon reasonable request to the authors

## Acknowledgements

We would like to thank Jake Horen for his support of this project. The work of Lora Henderson Smith was conducted with the support of the iTHRIV Scholars Program. The iTHRIV Scholars Program is supported in part by the National Center for Advancing Translational Sciences of the National Institutes of Health under Award Numbers UL1TR003015 and KL2TR003016 as well as by UVA. This content is solely the responsibility of the authors and does not necessarily represent the official views of NIH or UVA.

We do not have any conflicts of interest to report nor have we received any payments from a third party that could be perceived to influence, or give the appearance of potentially influencing, the submitted work.

## Notes

### Competing Interest Statement

The authors have declared no competing interest.

